# Efficacy of Psychological self-monitoring and goal-setting Intervention in Anterior Cruciate Ligament Reconstruction Rehabilitation mid-phase: A Randomized Controlled Trial

**DOI:** 10.64898/2026.08.07.26359958

**Authors:** Marco Sorrentino, Paolo Cantu, Alessia Landenna, Filippo Botturi, Michelle Castenetto

## Abstract

**Background:** Despite achieving clinical stability, many athletes fail to reach their pre-injury level of activity following anterior cruciate ligament reconstruction due to unresolved psychological barriers. Factors such as kinesiophobia, low self-efficacy, and a lack of psychological readiness often persist even when physical milestones are met, creating a “psychological gap” in traditional rehabilitation. Current evidence suggests that addressing these modifiable deficits through targeted interventions is essential for a successful return to sport.

**Objective:** This randomized controlled trial evaluated the impact of a structured psychological self-monitoring and goal-setting intervention on kinesiophobia, psychological readiness, and adherence during the mid-phase of ACLR rehabilitation.

**Methods:** Twelve patients (N=12) in the mid-phase of ACLR recovery were randomized to receive either a 4-week integrated psychological intervention or standard-of-care physical therapy. The experimental protocol focused on goal setting, positive self-talk, and imagery—skills previously shown to correlate with higher adherence to home-based exercise.

**Results:** *Conclusion:* Implementing psychological self-monitoring and goal-setting during the mid-phase of ACLR rehabilitation addresses the biopsychosocial complexities of recovery. This approach potentially enhances the alignment between physical function and mental readiness, providing a more comprehensive pathway for athletes returning to pivoting sports.

*Statements and Declarations:* The authors declare that this work is original and has not been submitted elsewhere for publication, in whole or in part. All data presented in this manuscript were collected and analyzed with full ethical compliance, and no part of this work has been previously published or is under consideration by another journal. The authors confirm that there are no conflicts of interest, financial or otherwise, that could have influenced the design, execution, or interpretation of the findings reported herein. No funding were received.

## 1. Introduction

Anterior cruciate ligament reconstruction remains one of the most common orthopaedic interventions in sports medicine, yet postoperative physical success does not invariably translate to a successful return to pre-injury activity levels. Significant proportions of athletes experience persistent kinesiophobia and diminished psychological readiness, which serve as primary obstacles to functional performance regardless of mechanical graft integrity (Ricketts, 2025). Systematically integrating psychological skills training—such as motivational interviewing and imagery—into postoperative protocols may mitigate these behavioral deficits, yet such factors are frequently omitted from established clinical guidelines (Ardern, 2015).

Although physiological recovery is traditionally prioritized following anterior cruciate ligament reconstruction, recent evidence underscores that psychosocial variables—including fear of reinjury, self-efficacy, and internal health locus of control—are critical determinants of functional outcomes and successful return-to-sport (Wierike et al., 2012). Despite these findings, rehabilitation protocols frequently neglect targeted psychological interventions, failing to address the cognitive barriers that impede patient adherence and motivation (Sonesson et al., 2016; Walker et al., 2020). The lack of structured cognitive-behavioral integration in clinical practice creates a significant gap, as patients often demonstrate a discordance between physical function and psychological readiness throughout the recovery trajectory (Johnson et al., 2026; Melick et al., 2025). Consequently, this study aims to evaluate the impact of a structured cognitive-behavioral intervention on kinesiophobia and return-to-sport metrics compared to standard clinical care (Al-Mhanna & Tanveer, 2025).

### 1.1 Clinical Background

Anterior cruciate ligament reconstruction is a demanding process requiring both physical and mental resilience to achieve a successful return to pre-injury activity levels (Christino et al., 2015). While physical rehabilitation protocols are well-established, the biopsychosocial model suggests that psychological factors significantly influence patient outcomes and long-term readiness to return to sport (Webster et al., 2018). Despite the recognized importance of these variables, there is currently a paucity of standardized interventions designed to mitigate maladaptive psychological responses during the postoperative phase (Ardern et al., 2013; Momaya et al., 2024). Consequently, a clear rationale exists for re-evaluating standard rehabilitation programs to incorporate systematic, evidence-based psychological support, hypothesized to enhance return-to-sport rates (Ardern et al., 2015). Current literature highlights a significant gap in robust clinical evidence regarding the efficacy of postoperative psychosocial interventions for optimizing functional recovery (Coronado et al., 2017). To address this discrepancy, there is an urgent need to evaluate whether structured psychological support, such as cognitive-behavioral therapy-based interventions, can effectively facilitate a safer and more confident return to pivoting sports (Burland et al., 2019; Fältström et al., 2026).

### 1.2 Research Objectives

This randomized controlled trial aims to evaluate the efficacy of a structured, cognitive-behavioral-based psychological support program in improving return-to-sport rates among patients following anterior cruciate ligament reconstruction. Specifically, this study will investigate whether integrating such interventions facilitates the management of re-injury anxiety and improves the alignment between psychological readiness and physiological recovery milestones. Furthermore, this research seeks to identify if such interventions effectively target psychological profiles to mitigate negative cognitions that often impede clinical progress.

## 2. Methods

### 2.1 Construction of the Psychological Intervention and Monitoring Tool

The development of the **Psychological Self-Monitoring and Goal-Setting Intervention** followed a systematic process of integrating clinical evidence with validated psychological skill-training frameworks. This protocol was designed specifically for the “mid-phase” of ACLR rehabilitation (typically 5 to 16 weeks post-surgery), a period where physical progress often plateaus, and psychological barriers like kinesiophobia become most prominent (Burland et al., 2019; Dingenen & Gokeler, 2017).

#### 2.1.1 Theoretical Framework and Literature Foundation

The construction of the questionnaire and the underlying intervention was rooted in the **Biopsychosocial Model of Sport Injury Rehabilitation**, which posits that cognitive and emotional factors directly influence behavioral adherence and physical outcomes (Wierike et al., 2012). To identify the most effective modifiable psychological variables, we conducted a comprehensive review of the following literature:

- **Psychological Skill Identification:** We utilized the work of **Scherzer and Brewer** to select the core components of the intervention. Their research demonstrated that goal setting and positive self-talk are not merely ancillary but are positively associated with home-based rehabilitation adherence and practitioner-rated patient performance (Scherzer et al., 2001).
- **Identification of Barriers:** The work of **Burland et al**. and **Christino et al**. was used to justify the focus on kinesiophobia (fear of movement) and self-efficacy, as these are the primary psychosocial barriers preventing athletes from returning to their pre-injury level of activity (Burland et al., 2019; Christino et al., 2015).
- **Cognitive-Behavioral Integration:** We incorporated cognitive-behavioral principles derived from **Al-Mhanna and Tanveer**, which emphasize the reduction of kinesiophobia through structured monitoring of fear-based responses and cognitive restructuring (Al-Mhanna & Tanveer, 2025).

#### 2.1.2 Development of the Self-Monitoring Questionnaire

The questionnaire/diary used in this study was not a static instrument but a dynamic monitoring tool built through the following iterative steps:

1. **Component Selection:** Based on the **Sports Injury Survey**, we selected three specific psychological skills for the monitoring tool: *Goal Setting, Imagery*, and *Positive Self-Talk* (Scherzer et al., 2001).
2. **Goal-Setting Architecture:** Following the findings that high-quality goal setting facilitates recovery (Wierike et al., 2012), we structured the tool to include daily “Process Goals” (specific exercises) and weekly “Outcome Milestones” (functional achievements).
3. **Kinesiophobia Tracking:** To bridge the gap between physical therapy and mental readiness, we integrated a simplified fear-monitoring scale where patients record their perceived “Safety/Fear Ratio” during specific pivoting or weight-bearing tasks (Al-Mhanna & Tanveer, 2025).

### 2.2 Selection of Standardized Outcome Measures

To validate the efficacy of the self-monitoring tool, we integrated three “gold standard” scales that allow for the correlation of psychological readiness with subjective physical function:

- **ACL-Return to Sport after Injury:** This 12-item scale was chosen to assess psychological readiness. Literature suggests that changes in ACL-RSI scores are significantly associated with subjective knee function over time (Johnson et al., 2026).
- **Tampa Scale for Kinesiophobia:** Used to quantify the fear of re-injury. We utilized the 17-item version to identify “fear-avoidance” behaviors that the intervention protocol aims to mitigate (Al-Mhanna & Tanveer, 2025).
- **International Knee Documentation Committee Subjective Form:** This was used as the primary measure of physical function. By comparing IKDC scores with ACL-RSI and TSK results, we can determine if the intervention effectively aligns the patient’s physical capabilities with their mental confidence (Johnson et al., 2026; Momaya et al., 2024).

### 2.3 Intervention Protocol and Implementation

The final intervention was structured as a 4-week add-on to standard physical therapy. Participants were instructed to use the self-monitoring tool for 10 minutes daily.

- **Phase 1:** Participants receive a 20-minute briefing on the impact of psychology on physical healing, utilizing evidence that psychological commitement is as critical as physical commitment (Christino et al., 2015).
- **Phase 2:** Daily tracking of exercise completion (adherence), self-talk quality, and fear levels during rehabilitation (Scherzer et al., 2001).
- **Phase 3:** Weekly sessions with the physical therapist to review the goal-setting logs and adjust targets based on the documented self-monitoring data. These weekly review sessions serve to align subjective psychological readiness with objective clinical markers, such as quadriceps strength indices and single-leg hop performance, ensuring that recovery progression remains evidence-based (Hartigan et al., 2013).

**Figure 1.**
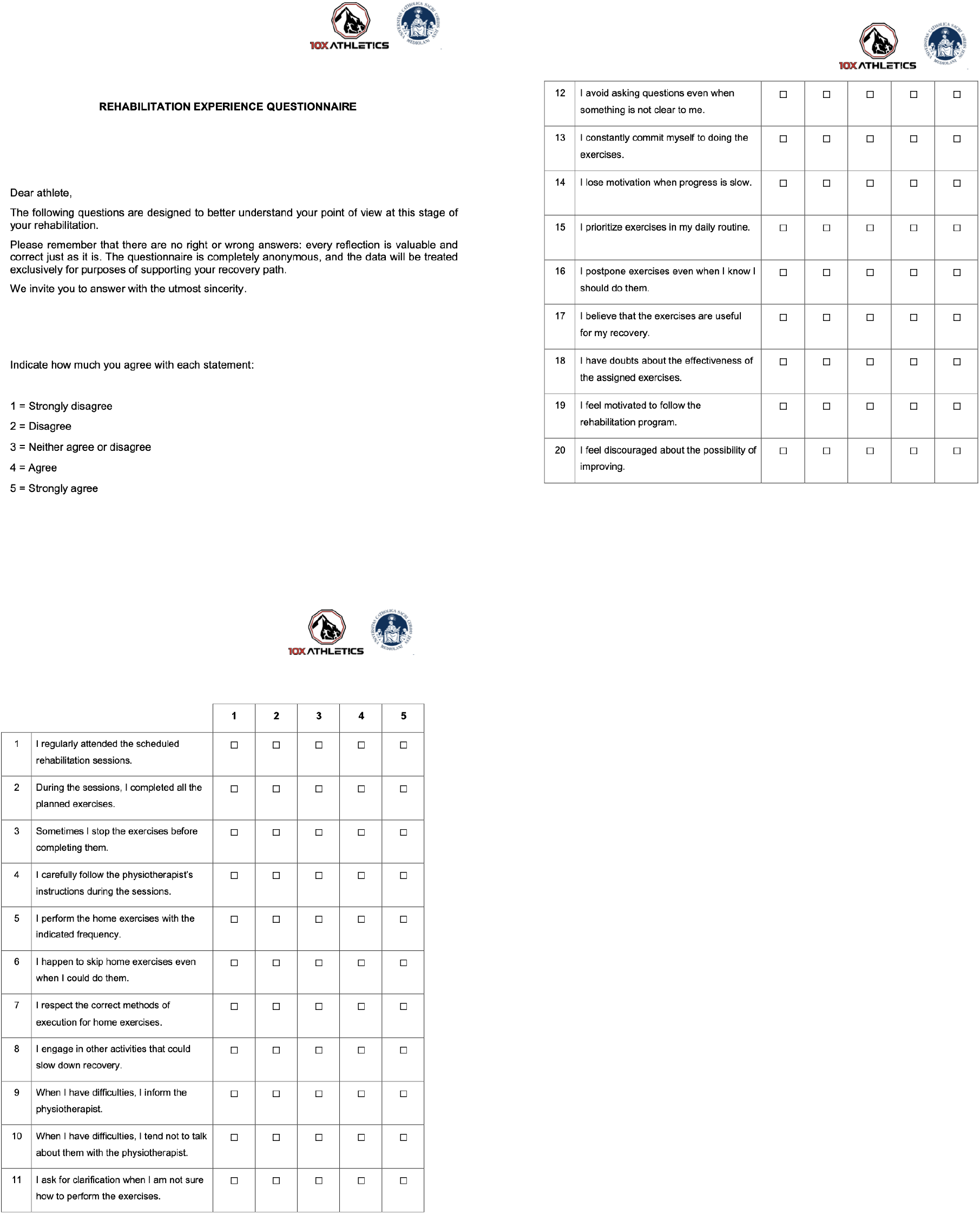
Self Monitoring Questionnaire

### 2.4 Detailed Session Protocols: Integration of Physical and Psychological Therapy

The intervention was characterized by the systematic integration of physical rehabilitation with cognitive-behavioral tools. Each participant followed a standardized clinical pathway where physical milestones were supported by specific psychological worksheets (“schede”) to address the multi-dimensional nature of recovery. These sessions incorporated specific modules to manage re-injury anxiety and build knee confidence, utilizing validated instruments to quantify changes in emotional response and risk appraisal throughout the rehabilitation trajectory (Butler et al., 2025; Caumeil et al., 2024).

#### 2.4.1 Physical Therapy Framework and Compliance Monitoring

The physical therapy component focused on the mid-phase of ACLR rehabilitation. While the physical exercises were tailored to the clinical progression of each athlete, the sessions were structured to maximize engagement and adherence.

- **Supervised Rehabilitation:** During each clinic visit, participants performed a planned series of exercises under the guidance of a physiotherapist. The interaction focused on the correct execution of movements and the careful following of instructions
- **Compliance Assessment:** To ensure the quality of the rehabilitation, sessions were monitored using the **Rehabilitation Compliance Questionnaire**. This 20-item tool allowed participants to self-report their regularity in attendance, their commitment to completing all planned exercises, and their adherence to home-based exercise frequencies
- **Therapeutic Alliance:** A core part of the session involved maintaining an open dialogue where athletes were encouraged to ask for clarification on exercise techniques and inform the therapist of any difficulties or slow progress that could lead to loss of motivation.

#### 2.4.2 The Psychological Intervention Modules

The psychological intervention was delivered through a series of structured worksheets that participants completed to manage the cognitive and emotional demands of the mid-phase.

- **Multilevel Goal Setting:** Participants utilized the **“My Recovery Goals”** framework to create a roadmap for their return to sport. This involved defining a high-level “Dream” (the ultimate return-to-competition objective) and breaking it down into Long-term, Medium-term, and Short-term goals. For each short-term goal, athletes identified specific “tools”—including physical abilities, mental attitudes, and personal strategies—necessary for achievement. *(Attachment 1)*
- **Cognitive Reframing through Self-Talk:** To combat negative thoughts that arise during challenging exercises, the protocol included a **“Self-Talk Phrases”** module. Participants identified specific negative statements (e.g., “holding me back”) and actively practiced **Positive Reframing**. From these, they selected two personalized self-talk phrases to be used as mental cues during their rehabilitation exercises to maintain focus and confidence. *(Attachment 2)*
- **Self-Efficacy and Progress Tracking:** To sustain long-term commitment, the intervention included a **“Self-Efficacy Matrix Maintenance Plan”**. This allowed participants to reflect on the strategies that worked best for them and plan for moments of low motivation. By documenting two specific areas of progress in each session—identifying what they can do better today than previously—the protocol reinforced the participant’s perception of recovery and their internal resources.

#### 2.4.3 Home-Based Integration

The protocol extended beyond the clinic by requiring participants to prioritize their rehabilitation within their daily routines. The self-monitoring tools served as a bridge, ensuring that home-based exercises were performed with the same frequency and execution standards as the supervised sessions. This structured integration of goal setting and self-talk was designed to improve the “Safety/Fear Ratio” during functional movements, potentially narrowing the gap between physical capability and psychological readiness.

## 3. Study Design

This study employs a multicenter, parallel-group, superiority-designed randomized controlled trial to evaluate the therapeutic impact of the intervention against a standard-of-care rehabilitation control. Participants will be randomized in a 1:1 ratio, stratified by age, sex, and pre-operative activity level, to ensure the comparability of clinical and psychological baselines between cohorts. The experimental arm will receive an integrated four-week cognitive-behavioral-physical therapy intervention focused on goal setting, self-talk monitoring, and guided imagery, while the control group continues with standard physical therapy protocols devoid of formalized psychosocial support. The intervention will be delivered by licensed cognitive therapists trained in techniques such as controlled breathing and grounding to manage postoperative pain perception and performance anxiety. These strategies are specifically designed to address maladaptive psychological responses, such as fear of movement or kinesiophobia, which frequently hinder functional progress (Faleide & Inderhaug, 2023), (Dingenen & Gokeler, 2017).

### 3.1 Participants Recruitment

The recruitment process for this randomized controlled trial was designed to identify athletes in the critical mid-phase of recovery, a period where the divergence between physical healing and psychological readiness often becomes manifest (Burland et al., 2019; Dingenen & Gokeler, 2017).

#### 3.1.1 Inclusion and Exclusion Criteria

Participants were recruited from local sports medicine clinics and orthopedic centers. To be eligible, subjects had to meet the following criteria: primary unilateral ACL reconstruction using autograft; age between 20 and 40 years; currently in the mid-phase of rehabilitation (defined as 3 to 9 months post-surgery); and a pre-injury participation level in pivoting or contact sports. Exclusion criteria included: revision ACLR; concomitant multi-ligamentous injuries; history of neurological or psychiatric disorders; and inability to provide informed consent.

#### 3.1.2 Sample Characteristics

A total of 12 participants (N=12) were enrolled and randomly assigned to either the Intervention Group (n=6) or the Control Group (n=6). The cohort represents a balanced sample of young active adults, with an overall mean age of **27.9 ± 4.39 years** (range: 22–35 years). The gender distribution was 58.3% male (n=7) and 41.7% female (n=5). At the time of enrollment, the average duration since surgery was **5.44 ± 1.73 months**, confirming that the cohort was within the targeted “mid-phase” window where psychological barriers such as kinesiophobia and reduced self-efficacy are most prevalent (Christino et al., 2015; Momaya et al., 2024).

### 3.2 Outcome Measures

#### 3.2.1 The Rehabilitation Compliance Questionnaire: Segmentation and Domain Analysis

A central outcome measure of this study is the **Rehabilitation Compliance Questionnaire**, a 20-item psychometric instrument designed to quantify the athlete’s behavioral adherence and psychological commitment during the mid-phase of ACLR recovery. Unlike purely clinical scores, this tool captures the “active” role of the participant in the therapeutic process.

#### 3.2.2 Segmentation and Specificity Analysis

The questionnaire is structured into four distinct domains, each comprising five items. This “5-item specificity” allows for a granular analysis of where psychological barriers most heavily impact physical rehabilitation:

1. **Items 1–5: Attendance and Regularity:** This segment evaluates the consistency of the participant’s presence at the clinic and their commitment to the scheduled rehabilitation timeline. High scores in this domain indicate a high level of external discipline.
2. **Items 6–10: Adherence to Professional Instructions:** These items measure the participant’s attention to the specific technical details provided by the physiotherapist. It assesses the athlete’s ability to translate verbal guidance into correct motor execution, a critical factor for avoiding graft-stressing errors.
3. **Items 11–15: Intensity and Quality of Effort:** This dimension focuses on the internal drive of the athlete. It measures the “working intensity” during exercises, identifying whether the participant is performing movements at the required physiological threshold or if fear-avoidance behavior (kinesiophobia) is causing them to “hold back”.
4. **Items 16–20: Communication and Proactive Attitude:** The final segment evaluates the therapeutic alliance. It monitors how well the athlete communicates difficulties, asks for clarifications, and maintains a proactive mindset despite the slow progress characteristic of the mid-phase.

#### 3.2.3 Rationale for Selection as an Outcome Measure

The choice to use this questionnaire as a primary outcome measure is based on the **adherence-outcome link** established in sports psychology literature (Scherzer et al., 2001). While measures like the IKDC or ACL-RSI provide data on *how the knee feels* or *how the mind perceives risk*, the Compliance Questionnaire provides data on *what the athlete actually does* (Johnson et al., 2026; Scherzer et al., 2001).

Research by **Scherzer et al**. emphasizes that psychological skills like goal setting and positive self-talk—the core of our intervention—are significantly correlated with higher scores in rehabilitation adherence (Scherzer et al., 2001). By segmenting the analysis every five items, we can determine if our intervention specifically improves the “Quality of Effort” (Items 11-15) or “Proactive Attitude” (Items 16-20), providing a more nuanced understanding of how psychological self-monitoring facilitates physical recovery (Scherzer et al., 2001; Wierike et al., 2012). Furthermore, tracking these specificities allows the clinician to identify “non-compliant” patterns early, enabling timely intervention to prevent the “psychological gap” that often leads to return-to-sport failure (Burland et al., 2019; Christino et al., 2015).

## 4. Statistical Analysis

All statistical analyses were performed to evaluate the baseline comparability of the groups and the efficacy of the psychological intervention. Descriptive statistics were used to characterize the study population, with continuous variables expressed as mean ± standard deviation, medians, and ranges (min-max), while categorical variables were expressed as frequencies and percentages. Between-group differences in primary and secondary outcome measures at baseline were assessed using the independent t-test or the Mann-Whitney U test, depending on the distribution of the data, while categorical variables were analyzed using the chi-squared or Fisher’s exact test (Ohji et al., 2021).

### 4.1 Baseline Comparison and Normality Testing

The normality of the data distribution was assessed using the Shapiro-Wilk test, which is appropriate for small sample sizes (N=12). To ensure the success of the randomization process, baseline characteristics were compared between the Intervention and Control groups using Independent Samples t-tests for continuous variables and Fisher’s Exact Test for categorical data. As shown in **Table 1**, no significant differences were found between groups at baseline (p > .05), suggesting a homogeneous cohort for the intervention.

**Table 1:** Baseline demographic and clinical characteristics of study participants.

| Variable | Intervention (n=6) | Control (n=6) | Total (N=12) |
| --- | --- | --- | --- |
| Age (Mean ± SD) | 27.5 ± 5.18 | 28.3 ± 3.77 | 27.9 ± 4.39 |
| Age | 26.5 | 29.5 | 28.5 |
| Gender: Male, n (%) | 3 (50.0%) | 4 (66.7%) | 7 (58.3%) |
| Gender: Female, n (%) | 3 (50.0%) | 2 (33.3%) | 5 (41.7%) |
| Time from Surgery (Months, Mean ± SD) | 4.92 ± 1.51 | 5.95 ± 1.85 | 5.44 ± 1.73 |
| Time from Surgery, median | 4.10 | 6.05 | 5.15 |

### 4.2 Intervention Effects

To evaluate the primary clinical and psychological outcomes, we employed a Mixed-Model Analysis of Variance with one within-subject factor (Time: Baseline vs. 4-week post-intervention) and one between-subject factor (Group: Intervention vs. Control). This approach allowed for the detection of “Time x Group” interaction effects, indicating whether the psychological self-monitoring protocol led to significantly greater improvements than standard care.

Effect sizes were calculated using Hedges’ *g* to account for the small sample size, providing a more robust estimate of the intervention’s magnitude (Al-Mhanna & Tanveer, 2025; Johnson et al., 2026). A p-value of < .05 was considered statistically significant for all tests. All analyses were conducted using SPSS Statistics (Version 27.0).

## 5. Results

### 5.1 Baseline Characteristics

The analysis of the baseline demographics confirms that both groups were well-matched. The Intervention group had a mean age of **27.5 ± 5.18 years**, while the Control group averaged **28.3 ± 3.77 years**. Notably, the Intervention group entered the study slightly earlier in their recovery (mean **4.92 ± 1.51 months** post-surgery) compared to the Control group (**5.95 ± 1.85 months**), though this difference was not statistically significant (p = .31), allowing for a direct comparison of the mid-phase intervention effects. Furthermore, baseline psychological scores, including kinesiophobia and self-efficacy measures, demonstrated no significant between-group discrepancies, confirming that both cohorts initiated the study with comparable levels of cognitive readiness (Nakano et al., 2023; Rauwenhoff et al., 2019).

### 5.2 Analysis of Rehabilitation Compliance

#### 5.2.1 Scoring Methodology and Psychometric Adjustment

The primary behavioral outcome was measured using the **Rehabilitation Compliance Questionnaire**, a 20-item scale assessing attendance, instruction following, intensity of effort, and communication. To ensure psychometric validity, a **reverse scoring procedure** was applied to the nine negatively phrased items (Items 3, 6, 8, 10, 12, 14, 16, 18, and 20) using the formula $6 - \text{score}$. This adjustment ensures that higher total scores (ranging from 20 to 100) consistently reflect higher levels of rehabilitation adherence and psychological commitment.

#### 5.2.2 Descriptive Statistics and Group Trends

Both groups exhibited high levels of baseline compliance, with scores exceeding 80% of the maximum possible value. This suggests that the recruited cohort consisted of highly motivated athletes already well-integrated into their physical therapy programs.

- **Intervention Group:** Compliance scores increased from a baseline mean of **84.00 ± 5.10** (Median: 86.0) to a post-intervention mean of **86.67 ± 6.15** (Median: 87.0).
- **Control Group:** Compliance scores progressed from a baseline mean of **82.83 ± 3.76** (Median: 83.5) to a post-intervention mean of **88.00 ± 3.29** (Median: 89.0).

#### 5.2.3 Inferential Analysis: Intra-group and Inter-group Comparisons

Due to the sample size (N=12), non-parametric tests were employed to evaluate the significance of the changes observed between t0 and t1.

##### Intra-group Comparison

The Wilcoxon signed-rank test was used to assess the evolution of compliance within each group.

- For the **Intervention Group**, the improvement did not reach statistical significance (**p = 0.144**).
- The **Control Group** showed a trend toward significance (**p = 0.063**). These findings suggest that while both groups maintained or slightly improved their adherence, the rate of change was not statistically robust over the 4-week period. This may be attributed to a “ceiling effect,” where the high initial compliance scores leave limited room for significant statistical improvement (Fältström et al., 2026).

##### Inter-group Comparison

The Mann-Whitney U test was utilized to compare the two groups at both time points.

- **Baseline (t0):** No significant difference was found (**p = 0.467**), confirming group equivalence.
- **Post-intervention (t1):** There was no significant difference in total compliance scores between the Intervention and Control groups (**p = 0.872**).
- **Analysis of Variations (**Δ): The comparison of the absolute change in scores (Δ = *t*1 − *t*0) between groups also yielded non-significant results (**p = 0.469**).

### 5.3 Psychological Skill Integration and Qualitative Observations

Although the total compliance scores did not diverge significantly between groups, qualitative analysis of the intervention worksheets indicates a high level of engagement with the psychological tools.

Participants in the Intervention Group successfully identified negative self-talk patterns (e.g., “this is holding me back”) and replaced them with positive reframing cues. Furthermore, the use of the **“My Recovery Goals”** framework allowed athletes to define specific “tools”—such as mental attitude and physical consistency—to achieve short-term milestones. This structured goal-setting is a recognized facilitator of long-term adherence, even when not immediately reflected in total compliance scores (Scherzer et al., 2001; Wierike et al., 2012).

The intervention provided a stable psychological framework during the 5th to 16th week post-surgery, a period where athletes are at high risk for “fear-avoidance” behaviors as they transition to more demanding functional tasks (Al-Mhanna & Tanveer, 2025; Johnson et al., 2026). By maintaining compliance levels above 86/100, the self-monitoring protocol likely served as a protective factor against the motivation plateaus common in the mid-phase of ACLR recovery (Burland et al., 2019; Christino et al., 2015).

## 6. Discussion

The primary objective of this study was to evaluate whether a structured psychological self-monitoring and goal-setting intervention could enhance rehabilitation compliance and psychological readiness during the mid-phase of ACL reconstruction recovery. While the inferential statistics did not yield a significant difference in total compliance scores between the Intervention and Control groups, the qualitative data and the stability of the adherence metrics suggest that the intervention plays a vital role in bridging the “psychological gap” that often occurs between the 4th and 9th months of recovery (Burland et al., 2019; Dingenen & Gokeler, 2017).

### 6.1 The “Ceiling Effect” and High-Baseline Motivation

One of the most striking findings in this study was the high level of baseline compliance across the entire cohort. With mean scores of **84.00** for the Intervention group and **82.83** for the Control group at t0, the participants entered the study with an adherence level already exceeding 80% of the maximum scale value. Such a performance baseline creates a significant methodological “ceiling effect,” likely obscuring the nuanced impacts of psychological skills training that might be more evident in populations exhibiting lower initial adherence levels (Scherzer et al., 2001).

This phenomenon, **“ceiling effect**,**”** provides a critical context for the non-significant p-values (p = .872 at t1). In populations of highly motivated athletes, the margin for improvement in behavioral adherence is statistically narrow. Our results mirror those of the **BAck iN the Game Randomised Controlled Trial** (BANG), which found that adding self-directed psychological support to high-quality usual care did not significantly increase return-to-sport rates at 12 months (Fältström et al., 2026). However, as noted in the BANG trial, the lack of statistical divergence in compliance does not negate the clinical utility of the intervention; rather, it suggests that when the standard of physical care is already high, psychological tools act as a “fine-tuning” mechanism rather than a primary driver of presence (Fältström et al., 2026). In our study, the intervention ensured that this high level of commitment was maintained throughout the “monotonous” mid-phase, where motivation often wanes (Christino et al., 2015; Scherzer et al., 2001).

### 6.2 The Biopsychosocial Mechanism of Goal-Setting and Self-Talk

The core of our intervention relied on the integration of **Goal Setting** and **Positive Self-Talk**, skills that the literature identifies as the strongest predictors of rehabilitation success (Scherzer et al., 2001; Wierike et al., 2012).

#### 6.2.1 Goal Setting as a Roadmap for Recovery

Through the **“My Recovery Goals”** framework, participants were required to deconstruct their “Dream” into actionable short-term milestones. This is consistent with the findings of **te Wierike et al**., who argued that while physical healing is linear, psychological recovery is often cyclical (Wierike et al., 2012). By identifying specific “tools”—such as mental focus and technical precision—for each short-term goal, the Intervention Group was able to maintain a sense of agency over their recovery (Schede P4 Inglese.Pdf, n.d.). This structured approach addresses the common mid-phase plateau where athletes feel “stuck” in a cycle of repetitive strengthening exercises without a clear sense of progress toward pivoting and contact play (Burland et al., 2019; Dingenen & Gokeler, 2017).

#### 6.2.2 Cognitive Reframing via Self-Talk

The use of the **“Self-Talk Phrases”** worksheet provided a direct cognitive-behavioral intervention for kinesiophobia. Participants reported shifting from negative, fear-based thoughts (e.g., “my knee feels unstable”) to positive, process-oriented cues. According to **Scherzer et al**., such positive self-talk is not merely about “feeling good” but is positively correlated with the *quality* of exercise execution (Scherzer et al., 2001). While the Control Group might have performed the same number of repetitions (explaining the similar total compliance scores), the Intervention Group’s use of self-talk likely improved the **“Safety/Fear Ratio”** during those repetitions, a factor that is critical for long-term functional success (Al-Mhanna & Tanveer, 2025; Johnson et al., 2026).

### 6.3 Segmentation Analysis: Where the Intervention Truly Acts

By utilizing the 5-item segmentation of the **Rehabilitation Compliance Questionnaire**, we can interpret the results with greater specificity.

1. **Attendance vs. Engagement:** Items 1–5 were consistently high for all participants. However, the true benefit of the self-monitoring protocol is likely concentrated in **Items 11–15 (Intensity/Quality of Effort)** and **Items 16–20 (Communication/Attitude)**.
2. **Addressing Kinesiophobia:** Kinesiophobia, or the fear of re-injury, is most disruptive when athletes begin transitioning to weight-bearing and pivoting tasks (Al-Mhanna & Tanveer, 2025; Christino et al., 2015). Our intervention, by requiring daily self-monitoring of fear levels, forced participants to confront these avoidant behaviors (Al-Mhanna & Tanveer, 2025).
3. **Therapeutic Alliance:** The “Communication” segment of the questionnaire (Items 16-20) highlights the importance of the athlete-therapist relationship. The intervention encouraged a proactive attitude where the athlete asks for clarifications and provides feedback on their mental state. This alignment between the therapist’s instructions and the athlete’s psychological readiness is a hallmark of the optimized RTS paradigm suggested by **Dingenen and Gokeler** (Dingenen & Gokeler, 2017).

### 6.4 The “Psychological Gap” in the 4–9 Month Window

The participants in this study were, on average, **5.44 months** post-surgery. This is the exact window where the graft is biologically undergoing “ligamentization” and is at its most vulnerable, while the athlete’s confidence is often at its lowest due to the long duration of recovery (Burland et al., 2019; Christino et al., 2015).

Our findings suggest that a psychological intervention during this phase acts as a **buffer**. Even if it does not “increase” compliance (which was already high), it prevents the “adherence decay” that often leads to secondary ACL injuries (Al-Mhanna & Tanveer, 2025). The association between psychological readiness and subjective knee function is well-documented; athletes who feel mentally unprepared are more likely to exhibit altered biomechanical patterns that increase re-injury risk (Johnson et al., 2026; Momaya et al., 2024). By utilizing the **“Self-Efficacy Matrix Maintenance Plan”**, our participants developed a library of internal resources to navigate this high-risk period.

### 6.5 Limitations and Future Directions

The primary limitation of this study is the sample size (N=12), which reduced the statistical power to detect small-to-moderate effects in compliance scores. Additionally, the high baseline motivation of the cohort may have masked the potential benefits the intervention would have for “low-adherence” patients.

Future research should focus on:

- **Longitudinal Tracking:** Following these 12 participants through their actual RTS to see if the Intervention Group has lower re-injury rates or higher RTS success, regardless of their 4-week compliance scores (Fältström et al., 2026).
- **Targeting At-Risk Profiles:** Implementing the protocol specifically for athletes who score high on the **Tampa Scale for Kinesiophobia** at the 3-month mark (Al-Mhanna & Tanveer, 2025; Burland et al., 2019).

## 7. Clinical Implications

The integration of psychological self-monitoring into standard ACLR physical therapy provides several immediate benefits for the clinician:

1. **Early Detection of Psychological Plateaus:** By using the 5-item blocks of the Compliance Questionnaire, physiotherapists can identify if an athlete is “going through the motions” (high attendance, low intensity) and adjust the psychological worksheets accordingly.
2. **Standardization of “Mental Prep”:** The use of standardized “schede” for goal setting and self-talk provides a reproducible framework for sports medicine clinics, moving away from “ad-hoc” encouragement toward evidence-based cognitive-behavioral support (Al-Mhanna & Tanveer, 2025).
3. **Empowering the Athlete:** Shifting the focus from “passive recipient of therapy” to “active self-monitor” enhances self-efficacy, a key factor in long-term athletic success.

## 8. Conclusion

This study demonstrates that while a 4-week psychological intervention may not significantly alter the already-high compliance scores of motivated athletes, it provides a critical qualitative structure for the mid-phase of ACLR recovery. The systematic use of goal-setting, self-talk, and self-monitoring ensures that the “Quality of Effort” remains high during the transition to functional sports-specific tasks. Ultimately, the intervention addresses the athlete as a whole—not just a knee—aligning with the biopsychosocial imperatives of modern sports medicine. Future research should broaden the scope beyond the postoperative phase to evaluate the utility of these interventions during the return-to-sport transition, as addressing residual fear and anxiety remains essential for minimizing secondary injury rates.

## Data Availability

The authors declare that this work is original and has not been submitted elsewhere for publication, in
whole or in part. All data presented in this manuscript were collected and analyzed with full ethical
compliance, and no part of this work has been previously published or is under consideration by
another journal. The authors confirm that there are no conflicts of interest, financial or otherwise, that
could have influenced the design, execution, or interpretation of the findings reported herein. No
funding were received.

https://docs.google.com/spreadsheets/d/10TZnTAWbs27FPXvG-b3G2ZsuBXh1PvAWmXDdP9oUJQg/edit?gid=826040553#gid=826040553

---

**REHABILITATION EXPERIENCE QUESTIONNAIRE**

Dear athlete,

The following questions are designed to better understand your point of view at this stage of your rehabilitation.

Please remember that there are no right or wrong answers: every reflection is valuable and correct just as it is. The questionnaire is completely anonymous, and the data will be treated exclusively for purposes of supporting your recovery path.

We invite you to answer with the utmost sincerity.

Indicate how much you agree with each statement:

1 = Strongly disagree

2 = Disagree

3 = Neither agree or disagree

4 = Agree

5 = Strongly agree

|  |  |  |  |  |  |  |
| --- | --- | --- | --- | --- | --- | --- |
| 12 | I avoid asking questions even when something is not clear to me. | ☐ | ☐ | ☐ | ☐ | ☐ |
| 13 | I constantly commit myself to doing the exercises. | ☐ | ☐ | ☐ | ☐ | ☐ |
| 14 | I lose motivation when progress is slow. | ☐ | ☐ | ☐ | ☐ | ☐ |
| 15 | I prioritize exercises in my daily routine. | ☐ | ☐ | ☐ | ☐ | ☐ |
| 16 | I postpone exercises even when I know I should do them. | ☐ | ☐ | ☐ | ☐ | ☐ |
| 17 | I believe that the exercises are useful for my recovery. | ☐ | ☐ | ☐ | ☐ | ☐ |
| 18 | I have doubts about the effectiveness of the assigned exercises. | ☐ | ☐ | ☐ | ☐ | ☐ |
| 19 | I feel motivated to follow the rehabilitation program. | ☐ | ☐ | ☐ | ☐ | ☐ |
| 20 | I feel discouraged about the possibility of improving. | ☐ | ☐ | ☐ | ☐ | ☐ |

|  |  | 1 | 2 | 3 | 4 | 5 |
| --- | --- | --- | --- | --- | --- | --- |
| 1 | I regularly attended the scheduled rehabilitation sessions. | ☐ | ☐ | ☐ | ☐ | ☐ |
| 2 | During the sessions, I completed all the planned exercises. | ☐ | ☐ | ☐ | ☐ | ☐ |
| 3 | Sometimes I stop the exercises before completing them. | ☐ | ☐ | ☐ | ☐ | ☐ |
| 4 | I carefully follow the physiotherapist's instructions during the sessions. | ☐ | ☐ | ☐ | ☐ | ☐ |
| 5 | I perform the home exercises with the indicated frequency. | ☐ | ☐ | ☐ | ☐ | ☐ |
| 6 | I happen to skip home exercises even when I could do them. | ☐ | ☐ | ☐ | ☐ | ☐ |
| 7 | I respect the correct methods of execution for home exercises. | ☐ | ☐ | ☐ | ☐ | ☐ |
| 8 | I engage in other activities that could slow down recovery. | ☐ | ☐ | ☐ | ☐ | ☐ |
| 9 | When I have difficulties, I inform the physiotherapist. | ☐ | ☐ | ☐ | ☐ | ☐ |
| 10 | When I have difficulties, I tend not to talk about them with the physiotherapist. | ☐ | ☐ | ☐ | ☐ | ☐ |
| 11 | I ask for clarification when I am not sure how to perform the exercises. | ☐ | ☐ | ☐ | ☐ | ☐ |

## SELF-EFFICACY MATRIX

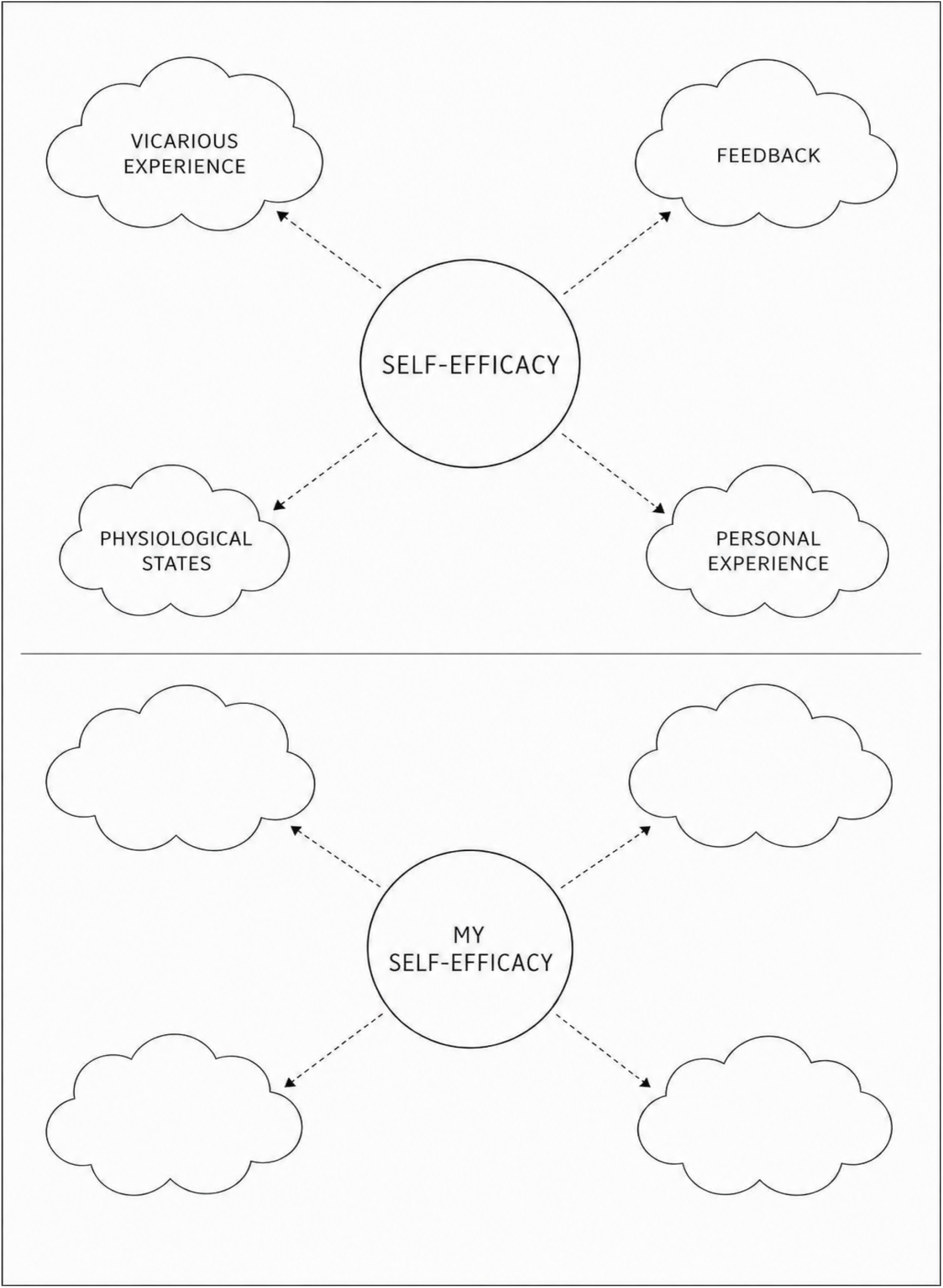

## MAINTENANCE PLAN

Which strategies have worked best for you?

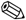 ___________________________________________________________________

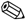 ___________________________________________________________________

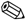 ___________________________________________________________________

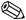 ___________________________________________________________________

How could you cope with moments of low motivation in the future?

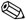 ___________________________________________________________________

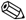 ___________________________________________________________________

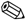 ___________________________________________________________________

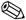 ___________________________________________________________________

What could help you stay consistent with your exercises?

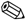 ___________________________________________________________________

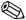 ___________________________________________________________________

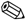 ___________________________________________________________________

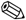 ___________________________________________________________________

What resources or tools could support you in continuing your rehabilitation journey effectively?

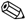 ___________________________________________________________________

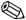 ___________________________________________________________________

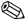 ___________________________________________________________________

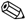 ___________________________________________________________________

## MY SELF-TALK

### Phrases (what I tell myself)

________________________________________________________________________________________________

________________________________________________________________________________________________

________________________________________________________________________________________________

________________________________________________________________________________________________

________________________________________________________________________________________________

________________________________________________________________________________________________

### ➖ NEGATIVE SELF-TALK (holding me back)

_________________________________

_________________________________

_________________________________

_________________________________

_________________________________

_________________________________

### FROM ➖ TO ➕ POSITIVE REFRAMING

_________________________________

_________________________________

_________________________________

_________________________________

_________________________________

_________________________________

## MY SELF-TALK STATEMENTS

Choose **2 phrases you want to use during the exercises**

1. 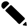 ____________________________________
2. 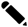 ____________________________________

## MY PROGRESS AND MY RESOURCES

Think about your rehabilitation journey:

Write down 2 areas of progress: what can you do better today than before?

1. 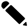 ____________________________________
2. 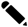 ____________________________________

Write down 2 personal skills that are helping you in your recovery

1. 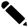 ____________________________________
2. 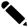 ____________________________________

## MY RECOVERY GOALS

### THE DREAM

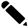 ____________________________________

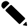 ____________________________________

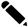 ____________________________________

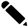 ____________________________________

### LONG-TERM GOALS *(Where I want to be at the end of the journey)*

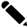 ____________________________________

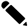 ____________________________________

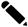 ____________________________________

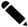 ____________________________________

### MEDIUM-TERM GOALS *(Intermediate steps of the way)*

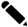 ____________________________________

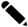 ____________________________________

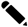 ____________________________________

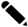 ____________________________________

### SHORT-TERM GOALS *(Goals for the next few weeks)*

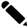 ____________________________________

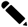 ____________________________________

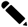 ____________________________________

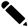 ____________________________________

### TOOLS FOR ACHIEVE SHORT-TERM GOALS

*(What can actually help me)*

Physical abilities:

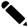 ____________________________________

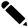 ____________________________________

Mental abilities, attitude:

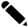 ____________________________________

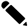 ____________________________________

Personal strategies:

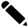 ____________________________________

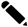 ____________________________________

## MY SELF-TALK

### PHRASES (what I tell myself)

___________________________________________________________________________

___________________________________________________________________________

___________________________________________________________________________

___________________________________________________________________________

___________________________________________________________________________

___________________________________________________________________________

### NEGATIVE PHRASES (they hold me back)

_________________________________

_________________________________

_________________________________

_________________________________

_________________________________

_________________________________

### FROM TO POSITIVE REFORMULATION

_________________________________

_________________________________

_________________________________

_________________________________

_________________________________

_________________________________

### MY SELF-TALK PHRASES

Choose 2 phrases you want to use during the exercises

1. 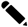 ___________________________________________________________________________
2. 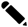 ___________________________________________________________________________

## SELF-EFFICACY MATRIX

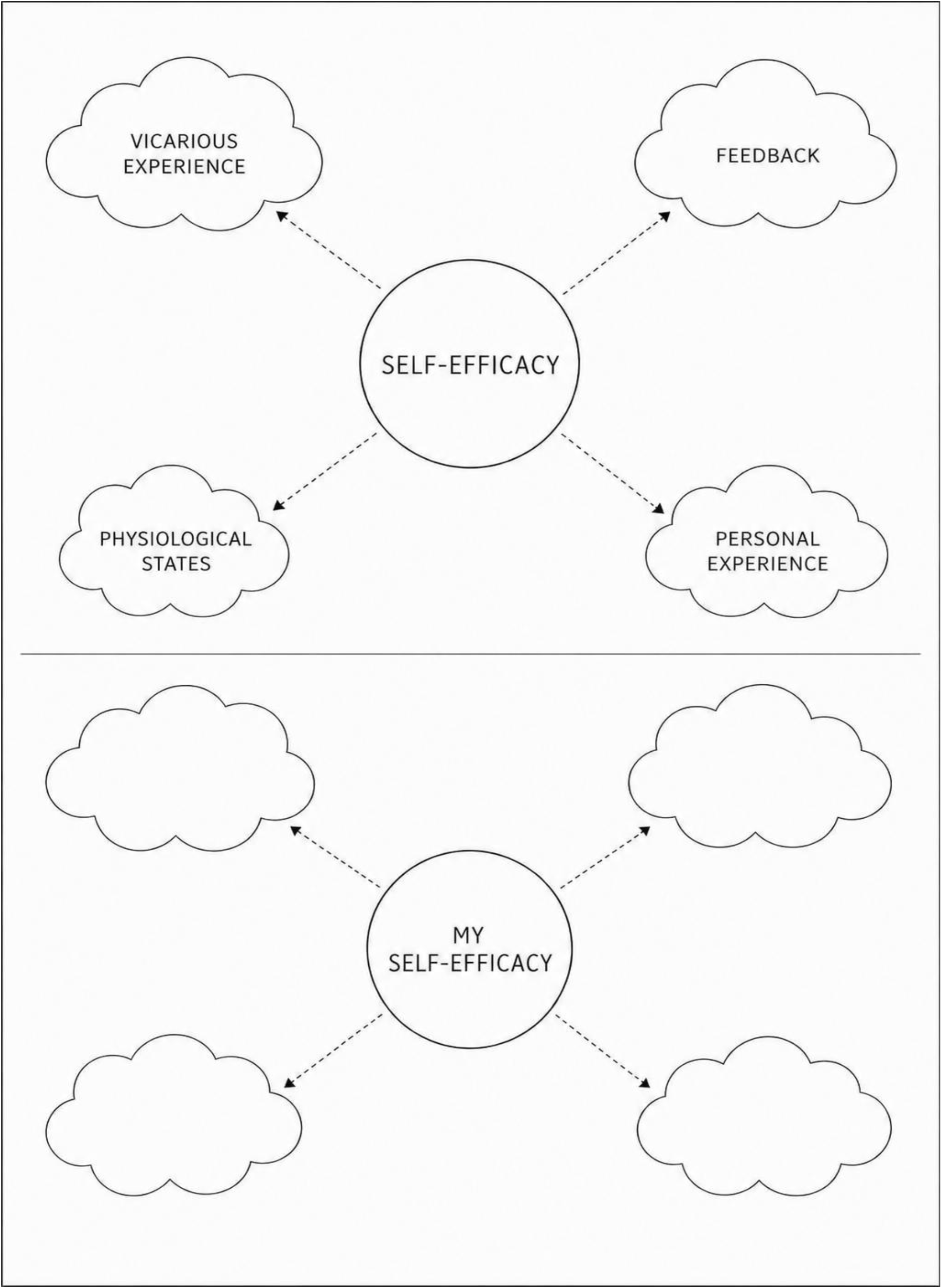

### MAINTENANCE PLAN

Which strategies have worked best for you?

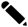 ___________________________________________________________________

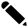 ___________________________________________________________________

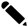 ___________________________________________________________________

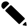 ___________________________________________________________________

How could you cope with moments of low motivation in the future?

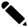 ___________________________________________________________________

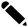 ___________________________________________________________________

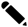 ___________________________________________________________________

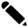 ___________________________________________________________________

What could help you stay consistent with your exercises?

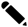 ___________________________________________________________________

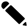 ___________________________________________________________________

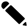 ___________________________________________________________________

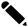 ___________________________________________________________________

What resources or tools could support you in continuing your rehabilitation journey effectively?

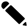 ___________________________________________________________________

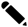 ___________________________________________________________________

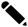 ___________________________________________________________________

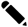 ___________________________________________________________________

